# Efficient multi-phenotype genome-wide analysis identifies genetic associations for unsupervised deep-learning-derived high-dimensional brain imaging phenotypes

**DOI:** 10.1101/2024.12.06.24318618

**Authors:** Bohong Guo, Ziqian Xie, Wei He, Sheikh Muhammad Saiful Islam, Assaf Gottlieb, Han Chen, Degui Zhi

## Abstract

Brain imaging is a high-content modality that offers dense insights into the structure and pathology of the brain. Existing genetic association studies of brain imaging, typically focusing on a number of individual image-derived phenotypes (IDPs), have successfully identified many genetic loci. Previously, we have created a 128-dimensional Unsupervised Deep learning derived Imaging Phenotypes (UDIPs), and identified multiple loci from single-phenotype genome-wide association studies (GWAS) for individual UDIP dimensions, using data from the UK Biobank (UKB). However, this approach may miss genetic associations where one single nucleotide polymorphism (SNP) is moderately associated with multiple UDIP dimensions. Here, we present Joint Analysis of multi-phenotype GWAS (JAGWAS), a new tool that can efficiently calculate multivariate association statistics using single-phenotype summary statistics for hundreds of phenotypes. When applied to UDIPs of T1 and T2 brain magnetic resonance imaging (MRI) on discovery and replication cohorts from the UKB, JAGWAS identified 195/168 independently replicated genomic loci for T1/T2, 6 times more than those from the single-phenotype GWAS. The replicated loci were mapped into 555/494 genes, and 217/188 genes overlapped with the expression quantitative trait loci (eQTL) of brain tissues. Gene enrichment analysis indicated that the genes mapped are closely related to neurobiological functions. Our results suggested that multi-phenotype GWAS is a powerful approach for genetic discovery using high-dimensional UDIPs.

## Introduction

Imaging genetics is an interdisciplinary field that seeks to unravel the intricate interplay between genetic variations and the phenotypes captured in imaging data to better understand the biological underpinnings of complex traits and diseases^1,2^. In particular, the emerging field of brain imaging genetics holds immense promise for advancing our understanding of the genetic basis of various neuropsychiatric disorders, cognitive functions, and behavioral traits^3^. To harness the potential of imaging genetics, researchers employ sophisticated methodologies and tools to generate imaging-derived phenotypes (IDPs), and conduct genetic association studies of these IDPs for investigating the relationships between genetic factors and neuroimaging measures^4–7^. The integration of genetic data, often obtained through techniques like single nucleotide polymorphism (SNP) arrays and sequencing technologies^8^, and the UK Biobank (UKB) is a rich prospective epidemiological study with a large-scale sequencing database^9^.

Additionally, UKB has a brain and body imaging extension, which aims to scan over 100,000 participants for the structural, diffusion and functional modalities^4^. Through genetic studies using large-scale, high-quality, and consistently acquired imaging data from the UKB, researchers identified genetic variants associated with specific IDPs and subsequently explored their implications for various neurological and psychological conditions^5,10–12^. A sophisticated image processing pipeline minimizes artifacts, standardizing images across various modalities and participants. Moreover, it produces a multitude of IDPs, offering distinct metrics of both brain structure and functions^13^. This approach has the potential to bridge the gap between the molecular basis and the macroscopic features of brain structure and function, shedding light on the mechanisms underlying both normal variations in brain traits and deviations that contribute to disease susceptibility^14^.

Besides traditional IDPs, our previous studies leveraged an unsupervised deep learning approach to derive robust, heritable and interpretable phenotypes, called Unsupervised Deep learning derived Imaging Phenotypes (UDIPs)^15^, from UKB T1 and T2 brain imaging data. We conducted single-phenotype genome-wide association studies (GWAS) and demonstrated that these specific phenotypic sets have higher individual heritability and can identify new loci not reported in the GWAS of traditional IDPs. The single-phenotype approach in GWAS has been routinely used in imaging genetics. Multiple IDPs are typically tested via GWAS one by one, and the results from individual GWAS are aggregated using multiple testing adjustments. However, such an approach does not exploit information contained in summary statistics from GWAS of related traits, especially for brain imaging IDPs, involving a large number of highly heritable brain morphological features. Previous GWAS have identified a great number of significant loci for individual IDP, by testing brain imaging phenotypes separately^12,16–18^. One of the largest GWAS of which, based on brain imaging data from 51,665 individuals, identified 199 significant loci^18^. Testing these related IDPs jointly could result in a further boost in statistical power to identify genetic variants with small effect sizes on individual IDPs^19^.

In addition, joint analysis of these traits is consistent with the notion that brain regions function as a synergistic unit and may leverage the discovery of genetic variants with distributed effects across regions and morphological measures^20^. It has been reported that large-scale multi-phenotype genomic analysis can elucidate the genetic architecture of shared components for complex diseases^21^. Several multi-phenotype GWAS approaches have been developed for genetic association analysis^19,22–25^. They were closely related to the multivariate analysis of variance (MANOVA)^26^, testing at a given SNP if the genotype is not associated with any of the phenotypes. MQFAM, also referred to as MV-PLINK, and MultiPhen, share a common approach involving multivariate regression^19,22^. Other multi-phenotype GWAS strategies, such as MultiABEL^24^, metaUSAT^25^ and MOSTest^19^, employ the MANOVA F-test or a chi-square test to compute the multivariate p-values. MTAG does not directly test the multivariate null hypothesis. Instead, it leverages association evidence from related traits to enhance genetic discoveries on a primary trait^27^. MultiABEL and metaUSAT operate on single-phenotype GWAS summary statistics to calculate the multivariate summary statistics. MOSTest does not take summary statistics, and instead it requires permutation on individual-level data. All these methods typically work efficiently for phenotypes with limited dimensions. For high-dimensional phenotypes, however, the lack of optimized software programs for a large number of phenotypes has become a bottleneck in the field. Here we introduce Joint Analysis of multi-phenotype GWAS (JAGWAS), designed to boost the power in brain imaging GWAS with hundreds of derived phenotypes for tens of thousands of possibly related individuals. JAGWAS takes single-phenotype GWAS summary statistics, estimates the correlation matrix from residualized phenotypes to improve computational efficiency, and computes the multivariate p-values analytically. Details of the JAGWAS procedure can be found in the Methods section.

We applied JAGWAS to deep-learning derived UDIPs to perform a 128-dimensional multi-phenotype GWAS. This analysis identified 6 times more loci than a Bonferroni-corrected analysis of 128 separate single-phenotype GWAS. The newly discovered loci are associated with various neurological disorders and critical brain morphology measurements, including cortical thickness, cortical surface area, and hippocampal subfield volume. The gene set and tissue enrichment analysis further linked these loci to neurobiological processes essential for cognitive function, emphasizing their potential relevance in neurological disorders like Alzheimer’s Disease (AD). By uncovering these connections, our study highlights the power of joint analysis of UDIPs in revealing complex genetic architectures and opens new pathways for understanding the biological underpinnings of AD, thereby aiding in the development of targeted therapies. This methodology showcases the potential to advance the genetic basis of neurological conditions through efficient multivariate statistical frameworks.

## Results

### Overview of the study

JAGWAS is a highly efficient summary statistics-based multi-phenotype GWAS method for up to hundreds of phenotypes (Methods: JAGWAS). In this study, we applied JAGWAS on 128 T1/T2 UDIPs building on our previous work^15^. The summary statistics for discovery, replication, and meta-analyses from these UDIPs were already available in the GWAS catalog^28^. These summary statistics, along with their estimated correlation matrices, were processed in JAGWAS to perform a multi-phenotype GWAS. The resulting summary statistics were fed into FUMA for locus clumping and functional gene mapping via the SNP2GENE function^29^. Independent genomic risk loci were identified, and a T-map was generated to visualize the brain regions affected by the top SNPs. Clustering analysis was performed using the optimal linear combination of the 128 UDIPs, with full methodological details outlined in the Methods: Clustering analysis. The mapped genes underwent further analysis in FUMA’s GENE2FUNC function to extract gene sets and tissue enrichment results for functional interpretation^29^. All the post-GWAS analyses focused exclusively on loci that had been successfully replicated in the independent replication cohort (Supplementary Data 1-2).

### Discovery and replication multi-phenotype GWAS

We performed JAGWAS on the 128 UDIPs using discovery (22,880 T1/T2) and replication cohorts (T1=12,359/T2=11,265). In the discovery stage, the genome-wide significance threshold was set at α = 5 × 10^−8^. JAGWAS produced much stronger signals compared to the minP approach, which selects the minimum p-value across single-phenotype GWAS for the 128 UDIPs, applying a Bonferroni-corrected threshold of 5 × 10^−8^ /128. The difference in p-value magnitude is particularly noticeable in Fig. 2a, with the Miami plot in Fig. 2b further illustrating the enhanced signals. JAGWAS identified 66,802/66,959 significant genetic variants clustering into 467/463 independent loci for T1/T2 (Supplementary Data 3). Using a 250 kb threshold, we merged the loci clumped by FUMA^29^ from the JAGWAS and minP results, and counted the number of merged loci contributed by JAGWAS and minP. Using the same approach, we compared our loci with those previously found by the big40 IDP GWAS^10^, which is one of the greatest efforts in identifying novel associations for brain imaging phenotypes. As can be seen in Fig. 2c, 108/113 loci previously identified by IDP GWAS were shared with JAGWAS on UDIPs. Additionally, all but one (both T1 and T2) loci discovered by the minP approach were also identified by JAGWAS. The total number of loci found by JAGWAS exceeded the combined total from both minP and IDP GWAS (218/233). Of all the loci identified, 384/376 were not detected by the minP approach, and 313/311 were exclusively identified by JAGWAS.

Replication analysis was carried out using an independent replication cohort, focusing on 66,802/66,959 significant variants identified during the discovery stage. Out of which, 21,422/19,357 significant variants, involving 195/168 independent loci for T1/T2, were successfully replicated at significance thresholds of 0.05/66,802 and 0.05/66,959 respectively (Supplementary Data 1).

### Meta-analysis

The discovery and replication cohorts were also meta-analyzed using a correlation matrix computed from the weighted average of covariance matrices from scaled residuals (discovery and replication). The meta-analysis results are consistent with the pooled analysis on the combined sample (discovery and replication), as shown in Supplementary Fig. 3. With a larger sample size, JAGWAS identified 156,650/145,708 variants which clustered into 844/776 independent association loci for T1/T2, at the genome-wide significance threshold of 5 × 10^−8^ (Supplementary Data 10). The Miami plot of JAGWAS vs. minP for meta-analysis can be found in Supplementary Fig. 3.

### Novel loci

As shown in Fig. 1c, JAGWAS identified hundreds of loci missed by the minP approach and previous IDP-GWAS. To better illustrate the loci uniquely found by our multi-phenotype GWAS method, a stringent approach in favor of minP was used. JAGWAS loci must pass a two-stage selection process (being significant in the discovery followed by replication) to ensure that any novel loci reported would not have been identified through minP in a meta-analysis, which combines the sample sizes of both the discovery and replication stages. With a larger sample size, the minP approach could find more loci, and still, 45/33 of the replicated loci from JAGWAS were not significant in minP results using a significance threshold of 5 ×10^-8^/128 (Supplementary Data 9). Among them, 9/4 loci identified by JAGWAS were still missed by the minP approach using a less stringent significance level of 5×10^-8^, without accounting for multiple testing of 128 phenotypes, as shown in Table 1. All the 9/4 loci in Table 1 were significant in both the discovery and replication cohorts of multivariate tests (p<0.05/66,802 = 7.48×10^-7^ and p<0.05/66,959=7.47×10^-7^, were considered significant in replication stage for T1/T2).

**Fig. 1.**
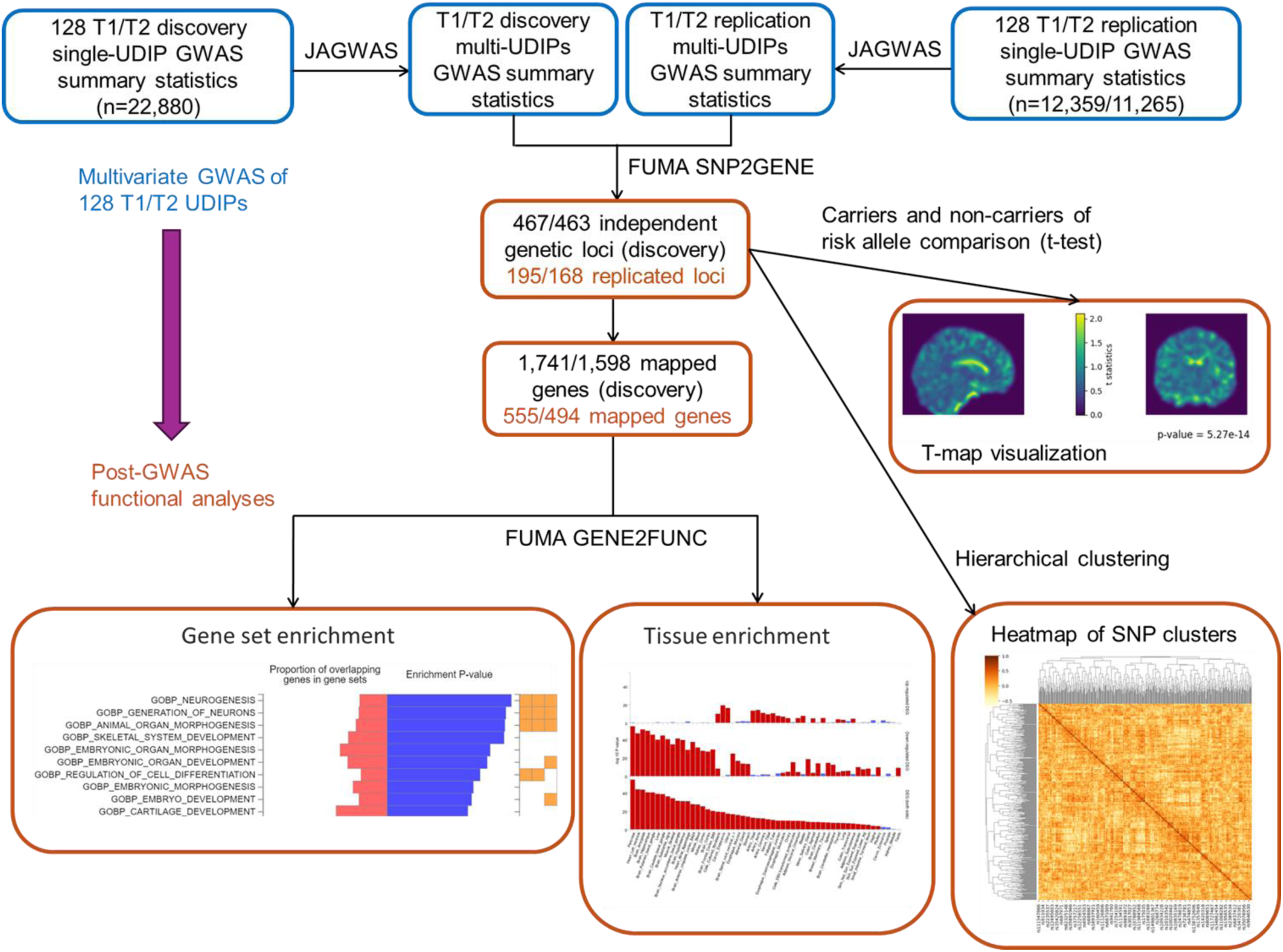
Overall pipeline of the study. The blue boxes represent the input and output of JAGWAS for the multi-phenotype GWAS of 128 T1/T2 UDIPs. The orange boxes represent the input and output from post-GWAS analyses using FUMA and clustering analysis.

**Fig. 2.**
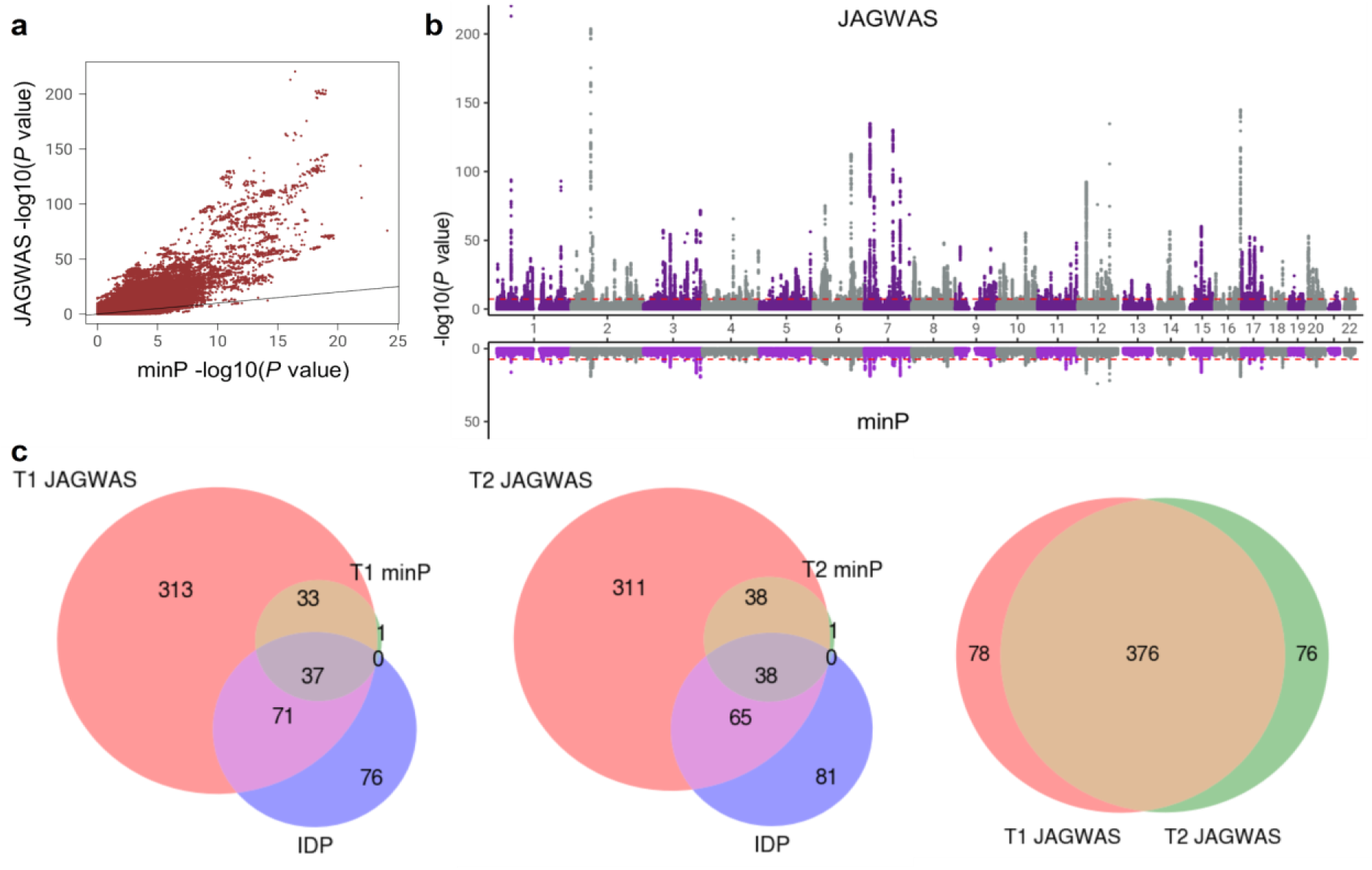
JAGWAS identifies new loci in the UKB (discovery cohort). **a**. Scatter plot of -log10 p-values from JAGWAS vs. minP (T1). **b**. Miami plot of -log10 p-values of JAGWAS vs. minP (T1), with JAGWAS on the top half and minP on the bottom half. The y axis represents the -log10 p-value and x axis shows the relative genomic location, grouped by chromosome, and the red dashed lines indicate the genome-wide significance threshold of 5 × 10^-8^. **c**. Venn diagrams displaying the number of loci identified by JAGWAS, overlapping between minP and previous IDP GWAS for T1 and T2, and overlapping between T1 and T2 UDIPs.

**Table 1.** JAGWAS identified novel loci previously missed by the minP approach. The table lists 9/4 (T1/T2) loci that were replicated in JAGWAS but missed by minP in a meta-analysis of discovery and replication, with an unadjusted threshold of 5×10^-8^. The p-values for discovery and replication cohorts of JAGWAS are listed, and the minP p-values are represented by P (minP meta).

| Lead SNP (T1) | CHR | POS | P (JAGWAS discovery) | P (JAGWAS replication) | P (minP meta) | Mapped genes | Neurological disorders/traits |
| --- | --- | --- | --- | --- | --- | --- | --- |
| rs708723 | 1 | 205739266 | 2.51x10<sup>-10</sup> | 3.34x10<sup>-7</sup> | 1.27x10<sup>-5</sup> | RAB29 | Parkinson's disease |
| rs745249 | 2 | 105460333 | 1.87x10<sup>-8</sup> | 3.87x10<sup>-7</sup> | 1.94x10<sup>-5</sup> | PANTR1 | Brain morphology (MOSTest), Cortical volume, Basal nucleus volume, Vertex-wise sulcal depth |
| rs1862133 | 5 | 173823888 | 6.96x10<sup>-15</sup> | 1.26x10<sup>-7</sup> | 3.51x10<sup>-7</sup> |  |  |
| rs11614730 | 12 | 79925802 | 9.33x10<sup>-22</sup> | 1.04x10<sup>-10</sup> | 6.38x10<sup>-7</sup> | SYT1<br>NOP56P3 | Attention deficit hyperactivity disorder or autism spectrum disorder or intelligence, Insomnia, General cognitive ability, Hippocampal volume |
| rs3008064 | 13 | 22998284 | 2.20x10<sup>-8</sup> | 2.39x10<sup>-7</sup> | 2.22x10<sup>-5</sup> |  |  |
| rs186347 | 14 | 59072226 | 1.14x10<sup>-30</sup> | 4.22x10<sup>-12</sup> | 4.20x10<sup>-7</sup> | RPL9P5<br>DACT1 | Language functional connectivity, Cortical volume, Hippocampal subfield right CA4 volume (head), Total PHF-tau |
| rs8039305 | 15 | 91422543 | 4.61x10<sup>-11</sup> | 4.64x10<sup>-7</sup> | 8.91x10<sup>-6</sup> | FURIN | Insomnia, Mental retardation, Bipolar disorder, Schizophrenia, Major depressive disorder, Ischemic stroke |
| rs35478105 | 16 | 77325325 | 1.39x10<sup>-13</sup> | 5.00x10<sup>-7</sup> | 1.90x10<sup>-5</sup> | ADAMTS18 | Insomnia, Alzheimer's disease, Cortical thickness, Cortical surface area |
| rs79973219 | 17 | 64040132 | 1.01x10<sup>-8</sup> | 2.47x10<sup>-7</sup> | 5.56x10<sup>-6</sup> |  |  |
| Lead SNP (T2) | CHR | POS | P (JAGWAS discovery) | P (JAGWAS replication) | P (minP meta) | Mapped genes | Neurological disorders/traits |
| rs1605141 | 5 | 90919961 | 4.66x10<sup>-18</sup> | 2.46x10<sup>-8</sup> | 3.13x10<sup>-4</sup> |  |  |
| rs186347 | 14 | 59072226 | 1.55x10<sup>-20</sup> | 1.14x10<sup>-8</sup> | 1.17x10<sup>-2</sup> | RPL9P5<br>DACT1 | Language functional connectivity, Cortical volume, Hippocampal subfield right CA4 volume (head), Total PHF-tau |
| rs117068593 | 14 | 93118229 | 2.42x10<sup>-14</sup> | 8.83x10<sup>-12</sup> | 1.16x10<sup>-4</sup> | RIN3 | Alzheimer's disease, Subcortical volume (MOSTest), Vertex-wise sulcal depth |
| rs6097618 | 20 | 52448936 | 9.44x10<sup>-14</sup> | 2.54x10<sup>-7</sup> | 3.56x10<sup>-3</sup> | RNU7-14P,<br>SUMO1P1 | Brain region volumes, Cortical volume, Vertex-wise cortical thickness, Vertex-wise cortical surface area |

**Table 2.**
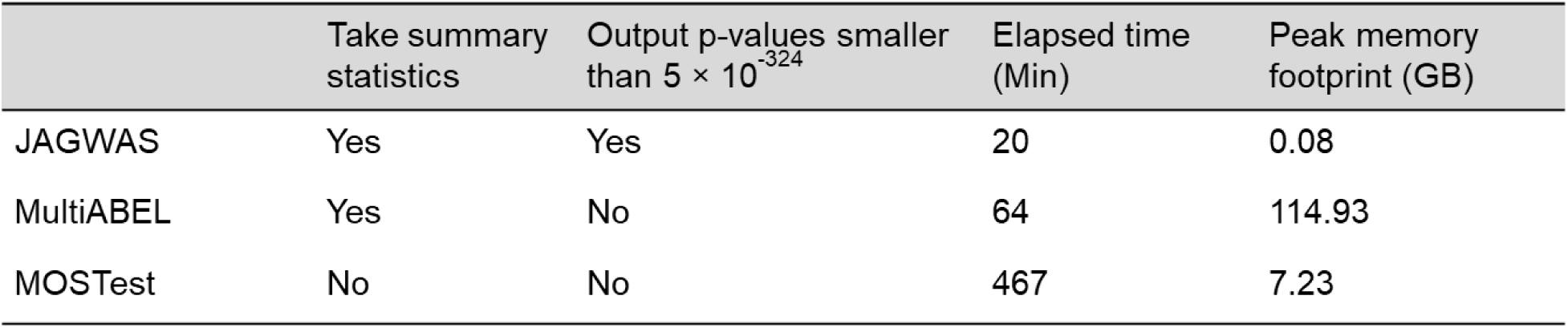
Features of JAGWAS and other multi-phenotype GWAS methods.

| | Take summary statistics | Output p-values smaller than $5 \times 10^{-324}$ | Elapsed time (Min) | Peak memory footprint (GB) |
| --- | --- | --- | --- | --- |
| JAGWAS | Yes | Yes | 20 | 0.08 |
| MultiABEL | Yes | No | 64 | 114.93 |
| MOSTest | No | No | 467 | 7.23 |

Many interesting associations were found (Table 1). For example, rs708723 (p = 2.51×10^-10^, T1 UDIPs) on chromosome 1 was reported to be associated with Parkinson’s disease (PD)^30^. Other loci associated with neurological disorders were: rs8039305 (p = 4.61×10^-11^, T1 UDIPs) on chromosome 15 was identified by MTAG for associations with bipolar disorder, schizophrenia, and major depressive disorder^31^; the shared loci rs186347 (p = 1.14×10^-30^/1.55×10^-20^, T1/T2 UDIPs) was associated with language functional connectivity^32^; rs35478105 (p = 2.51×10^-10^, T1 UDIPs) on chromosome 16 was associated with insomnia^33^. A total of 7 of these loci showed associations with brain measurement or cortical measurement. Examples are rs11614730 of T1 on chromosome 12 (p = 9.33 ×10^-22^, T1 UDIPs) was reported to be associated with cortical thickness by MOSTest^19^; rs6097618 (p = 9.44 ×10^-14^, T2 UDIPs) was associated with brain region volumes, cortical volume, vertex-wise cortical thickness, and vertex-wise cortical surface area^12,34,35^; the shared loci rs186347 was associated with cortical volume and hippocampal subfield right CA4 volume^34,36^, hippocampal subfield volume was reported to be closely associated with verbal memory after first-ever ischemic stroke^37^.

### Enrichment of neurobiologically relevant genes and differentially expressed genes

Through FUMA^29^, 555/494 genes were mapped respectively for T1/T2 replicated loci, and 217/188 of them overlapped with expression quantitative trait loci (eQTL) of one or more of the 13 Genotype-Tissue Expression (GTEx) v8 brain tissues (Supplementary Data 4). We used FUMA SNP2GENE function to perform gene set and tissue enrichment analysis, which are based on the hypergeometric distribution test with a lower p-value indicating greater over- representation. Tissue specificity is tested using the differentially expressed gene (DEG) sets defined for each of the GTEx v8 tissue types. Using T1 as an example, the gene sets demonstrate significant enrichments for biological processes, including neurogenesis (p = 3.54 × 10^-8^), generation of neurons (p = 5.24 × 10^-8^), animal organ morphogenesis (p = 1.52 × 10^-8^), and regulation of neuron differentiation (p = 5.49 × 10^-7^).

Through gene set enrichment analyses, we identified 19 significant biological process ontology sets for T1 UDIPs, many of which related to neuronal development and differentiation, with the top 10 listed in Fig. 3a (Supplementary Data 5). The tissue enrichment results of JAGWAS and minP classified by organ systems for T1 are listed in Fig. 3b (Supplementary Data 6). The most enriched tissues from JAGWAS are pancreas (p = 1.44 × 10^-10^), putamen basal ganglia (p = 3.51 × 10^-9^) and most of the brain tissues categorized under the nervous system. The top 5 enriched brain tissues are putamen basal ganglia (p = 3.51 × 10^-9^), amygdala (p = 7.22 × 10^-9^), caudate basal ganglia (p = 1.65 × 10^-8^), substantia nigra (p = 1.74 × 10^-7^) and hypothalamus (p = 3.24 × 10^-7^). In contrast, none of the tissues displayed statistically significant enrichment for minP results. The corresponding Fig. a-c for T2 are in Supplementary Fig. 4.

**Fig. 3.**
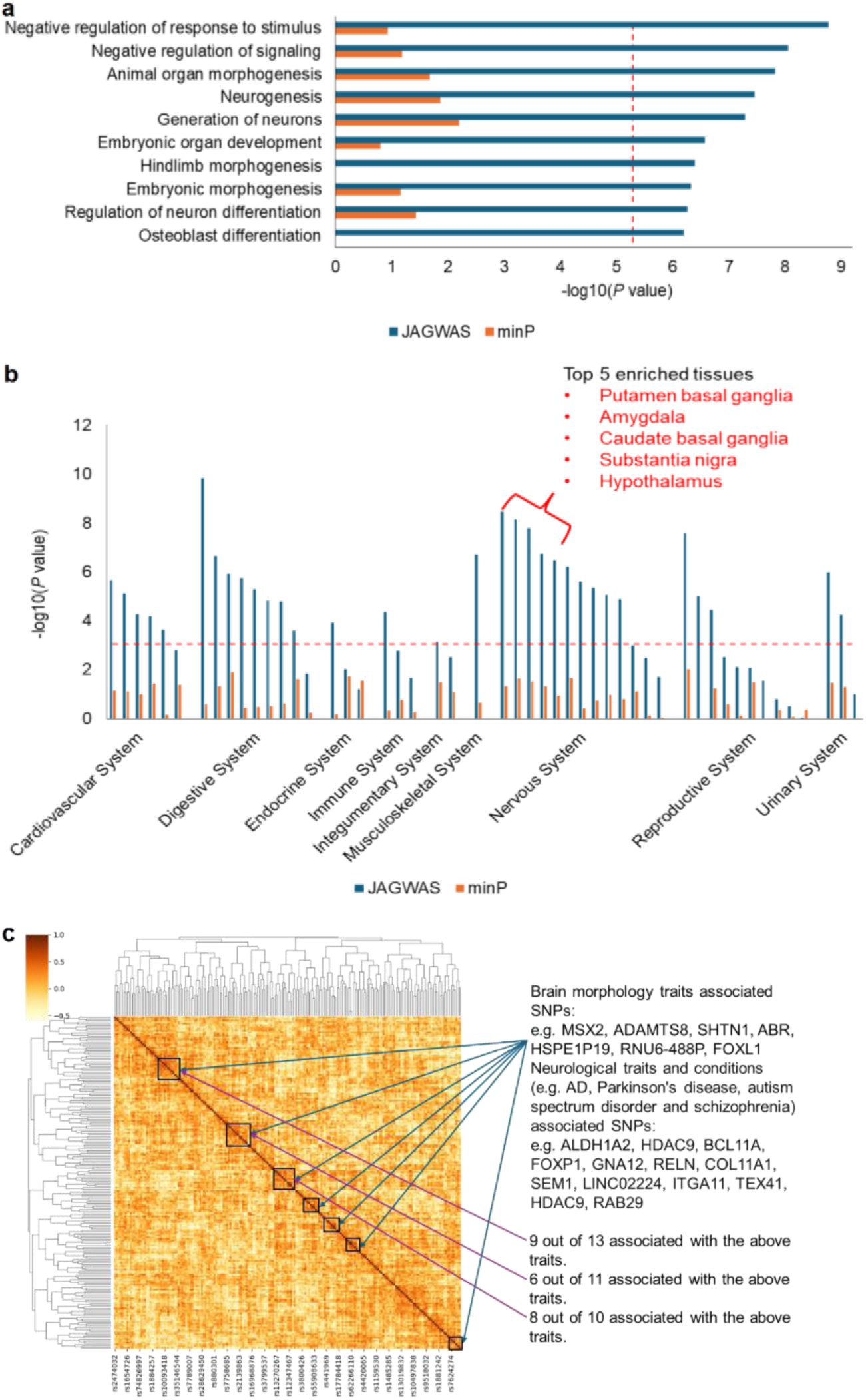
Enrichment and clustering analyses indicate high neurobiological relevance of JAGWAS hits (T1). **a.** Results from gene-set enrichment analyses for JAGWAS and minP. The top 10 most significant Biological Process Gene Ontology sets for JAGWAS (blue bars) are shown on the y axis, - log10 p-value on the x axis. The corresponding minP p-values are represented by the orange bars. The red dashed significance line (0.05/7,750) has been adjusted using a Bonferroni correction. **b.** Results from tissue enrichment analyses for JAGWAS (blue bars) and minP (orange bars). The GTEx v8 54 tissue types classified by organ systems are shown on the x axis, -log10 two-sided DEG p-value on the y axis. The red dashed significance line (0.05/54) has been adjusted using a Bonferroni correction. **c.** Clustering lead SNPs by their vectors of optimal weights on the 128 UDIPs to group SNPs with similar pleiotropic effects. Darker colors indicate highly similar SNPs based on cosine similarity.

### Clustering reveals lead SNPs with similar pleiotropic effects

Clustering lead SNPs based on their best linear combination of UDIPs (Supplementary Data 7) identifies clusters with a great proportion of SNPs associated with brain morphology traits and neurological traits for T1, see Fig. 3c. Specifically, we perform hierarchical clustering based on the cosine similarity (Supplementary Data 8) between the vectors of optimal weights for each lead SNP (Methods: Clustering analysis). See supplementary information for details of the derivation of the weights. The color in the plot indicates cosine similarity, with darker colors representing highly similar SNPs. We analyzed the SNP clusters given by performing hierarchical clustering on the UDIP weights. A high proportion of SNPs within the clusters are associated with brain morphology traits and neurological traits, see Fig. 3c. For example, recent articles pointed out that *HDAC9*- mediated calmodulin deacetylation induces memory impairment in AD, and *FOXP1* plays a crucial role in the molecular mechanisms underlying schizophrenia^38,39^; *GNA12* is associated with Parkinson’s disease and bipolar disorder^40,41^.

### Comparison between JAGWAS and other tools

We compared JAGWAS with other multi-phenotype GWAS methods, including MultiABEL and MOSTest, in addition to minP. MultiABEL takes single-phenotype GWAS summary statistics and employs Pillai’s trace MANOVA to calculate the multivariate p-value from an F-statistic, which in large samples is close to the chi-square test utilized by JAGWAS. In contrast, MOSTest requires access to individual-level data and generates single-phenotype GWAS results by permuting genotype vectors for each SNP and then integrates effects across the phenotypes using the Mahalanobis norm to produce a multivariate summary statistic. Features of these multi-phenotype GWAS methods are presented in Table. 2. All the analyses were running on an x86 64-bit Linux system with 48 cores. The elapsed time and peak resident memory size (RES) were recorded for each method. We benchmarked JAGWAS, MOSTest and MultiABEL by conducting a multi-phenotype GWAS with roughly 8.1 million SNPs. We found that JAGWAS spent only 32.3% of the time and 0.07% of peak memory compared to MultiABEL. This efficiency is attributed to JAGWAS’s optimized C++ implementation and memory-efficient file handling. Another advantage of JAGWAS is that it provides an option to output p-values on the log scale, overcoming the double precision limit of MultiABEL for extremely significant results, as shown in Supplementary Fig. 5.

Starting from summary statistics and using a flexible input/output (I/O) strategy, JAGWAS was able to obtain the results faster while using less memory than MOSTest. Lastly, both JAGWAS and MultiABEL can account for sample relatedness by using the single-phenotype GWAS summary statistics from mixed effects models. All three methods produced consistent results with each other, as in Supplementary Fig. 5-6, which is assuring.

### SNP effect visualization

We visualized the effect of each replicated lead SNP by comparing the MRI imaging data between homozygous carriers and non-carriers (Method: Visualizing SNP effect). Interestingly, we found many SNPs with localized patterns of difference that had not been identified previously, suggesting that genetic variations at these loci could be driving localized brain changes and highlighting the power of our method. For example, rs11706279 displayed a significant pattern of differences in the lingual gyrus and occipital lobe, regions crucial for visual processing and memory^42,43^. No prior associations have been documented for this SNP in the GWAS Catalog or the UK Biobank big40 PheWeb^10,28,44^. Research suggested that in AD patients with depression, functional connectivity between the dorsal anterior cingulate cortex and the right occipital lobe and right lingual gyrus is reduced^45^. Occipital lobe is also involved in schizophrenia, but the exact nature of the relationship is not fully understood^46^. This SNP is located in an intron of *FOXP1*, which has been associated with both Alzheimer’s disease and schizophrenia ^39,47^.

## Discussion

Our results showed that applying the multivariate approach JAGWAS to UDIPs greatly boosted the power of loci discovery for brain imaging endophenotype GWAS. We discovered 6 times more loci than minP, and two times more loci than the previous IDP GWAS that used a similar or larger sample size^5,10^. With JAGWAS, we identified a large number of loci that were missed by the minP method, which indicated the high level of pleiotropy of the variants associated with the high-dimensional UDIPs. Through post-GWAS functional analyses, we found that our replicated loci showed significant enrichment in associations with brain and cortical morphological traits, as well as neurological disorders, attesting to the power of JAGWAS in uncovering new loci.

We showed that the loci and genes found by JAGWAS have high neurobiological relevance. First of all, genes associated with biological processes like neurogenesis, generation of neurons, and regulation of neuron differentiation are enriched within the gene set; all of these processes play an essential role in forming the nervous system. Neurogenesis primarily occurs during embryonic development^48^, which corresponds with the embryonic development terms in Fig. 3a. However, neurogenesis also continues in certain regions of the adult brain, notably in the hippocampus, which is associated with memory and learning^48^. Generation of Neurons involves the differentiation of neural stem cells into neurons, followed by their migration to the appropriate location in the brain, and then integration into existing neural networks^49^. Neuron differentiation is a complex process that is regulated by a variety of factors^50^. These processes are essential for brain plasticity, the ability of the brain to adapt and reorganize itself, especially in response to learning, experience, and injury. None of the top 10 significant biological processes were over-represented in minP genes. This might indicate that multivariate approaches like JAGWAS can capture biological pathways of the brain missed by the single-phenotype approaches.

In addition, tissue enrichment results from FUMA indicated the genes have a great enrichment in all tissues from the nervous system, with the top five enriched tissues for T1/T2 being amygdala, hypothalamus, and three tissues come from basal ganglia: putamen, caudate and substantia nigra. Amygdala and putamen are actively involved in memory formation, learning and cognitive functioning^51,52^. Hypothalamus, long recognized for its role in nutrient sensing and the regulation of arousal and motivation, is also actively involved in updating both associative and non-associative memories^53^. It has been reported that there could be strongly reduced volumes of putamen and thalamus in AD^52^. Caudate and substantia nigra are implicated in movement control; and notably, substantia nigra was affected in Parkinson’s disease^54^. In contrast, all the brain tissues have non-significant enrichment for minP of T1/T2, which further highlights the great power of JAGWAS in uncovering the genetic architecture of the brain (Fig.3 and supplementary Fig. 4).

An additional benefit of UDIPs is that we can cluster SNPs into groups with similar pleiotropic effects (Supplementary Data 8). The clusters identified in Fig. 3c show a high proportion of SNPs that are associated with brain morphology and neurological traits, including AD and PD. Specifically, the most enriched clusters contain 9 out of 13, 6 out of 11, and 8 out of 10 SNPs linked to these traits. SNPs within these clusters exhibit high cosine similarity based on their optimal weights across the UDIPs, with several SNPs associated with the same neurological traits. A notable example includes three neighboring SNPs in the cluster map: rs12705150 (*RELN*, p = 2.16 × 10^-27^), rs11614730 (*SYT1*, p = 1.84 × 10^-38^) and rs12722976 (*COL11A1*, p = 2.53 × 10^-33^), with cosine similarities of 0.51, 0.39, and 0.24 between each other. *RELN* has been linked to neuropathology in AD^55^, while *COL11A1* is associated with AD polygenic risk score^56^; additionally, *SYT1* and *COL11A1* have been reported to be involved in conditions such as attention deficit hyperactivity disorder (ADHD), autism spectrum disorder (ASD), or intelligence, indicating pleiotropy^57^. Another example of three adjacent SNPs are: rs8039305 (*FURIN*, p = 1.13 × 10^-23^), rs4775006 (*ALDH1A2*, p = 5.13 × 10^-103^) and rs12256551 (*MIR1915HG*, p = 2.55 × 10^-58^), with cosine similarities of 0.48, 0.23, and 0.15 between them. Both *FURIN* and *MIR1915HG* have been linked to insomnia, while *ALDH1A2* and *FURIN* are associated with schizophrenia^58,59^. Furthermore, *ALDH1A2* has been reported in association with both AD and cognitive decline in AD^60,61^. These results suggest that the clustering analysis effectively captured the functional similarity of the lead SNPs based on their optimal weights across the 128 UDIPs. The observed enrichment and clustering findings align with our novel loci discoveries, both *FURIN* and *SYT1* are present in Table 1. Neurological disorders such as AD and PD are strongly linked to abnormal brain morphology and structural measures, which are influenced by differential gene expression in specific brain regions^62^. We further looked into these replicated lead SNPs by comparing the MRI imaging data between homozygous carriers and non-carriers (Fig. 4 and Supplementary Fig. 7). Localized patterns of difference were found in brain regions like lingual gyrus, occipital lobe and insular cortex, while no such association was previously found. Our multivariate approach is tailored to reveal this kind of relationship beyond the single-phenotype approach.

**Fig. 4.**
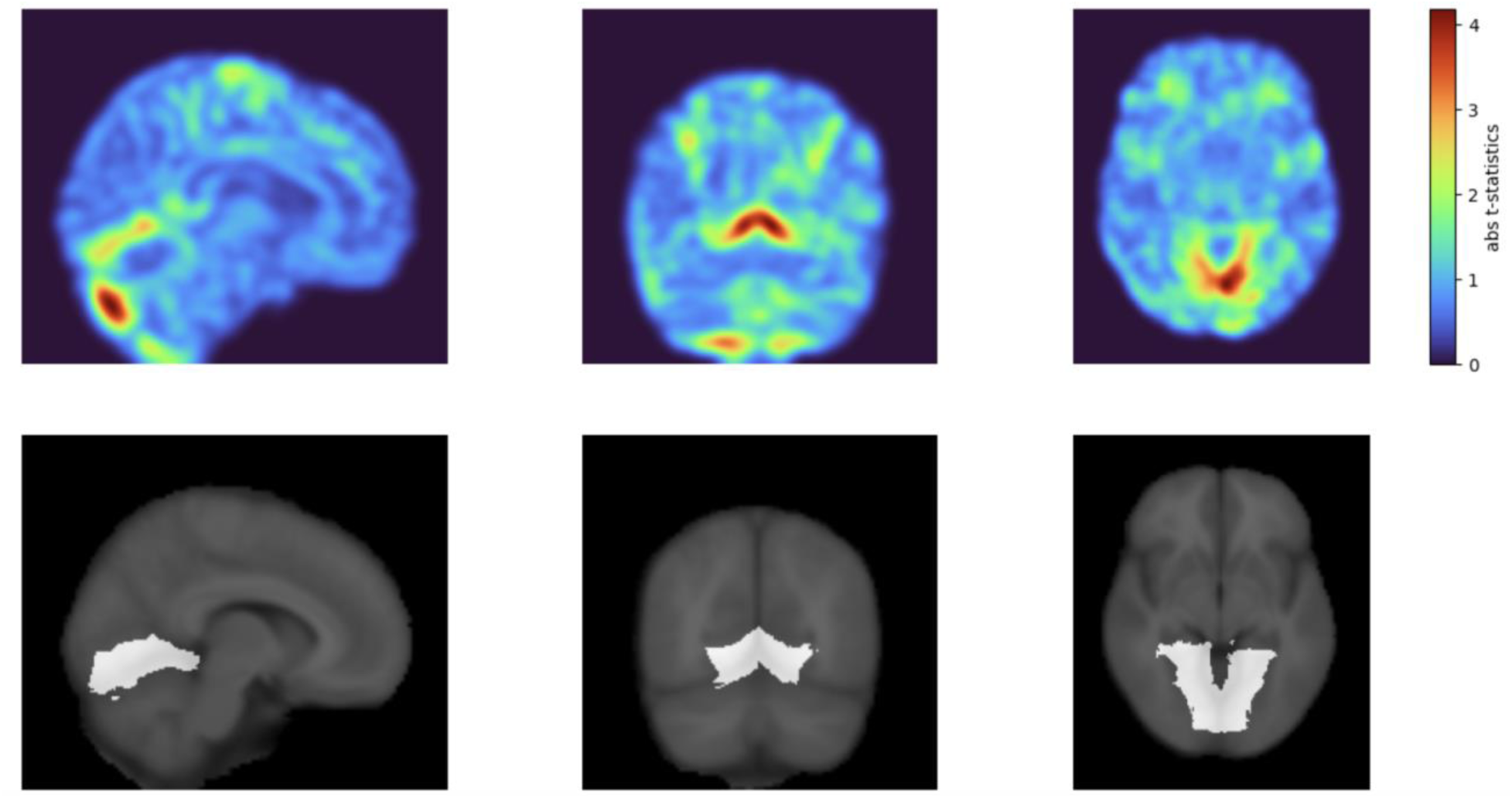
Visualization of the SNP effect of rs11706279. The top figure shows the voxel-wise absolute t-statistics for the contrast between homozygous carriers and non-carriers. The bottom figures highlight the lingual gyrus.

JAGWAS was designed to jointly analyze GWAS summary statistics of hundreds or even thousands of phenotypes, with exceptional computational efficiency. We compared the feature of JAGWAS to several commonly used multi-phenotype GWAS tools and did a detailed benchmark test. The key features of JAGWAS include: 1) the ultra-efficiency in time and memory usage through an optimized C++ implementation; it can analyze 128 summary statistics files of 8.1 million SNPs in approximately 20 minutes (Table. 2). 2) JAGWAS supports various summary statistics formats, including score tests (score and variance), Wald tests (beta and standard errors), or simply z-scores. 3) JAGWAS can output -log10 p-value to get p-values smaller than the double precision limit. This leverages the large-scale meta-analyses and high-dimensional multi-phenotype GWAS by allowing the output of extremely significant p-values.

With the large gain of power of using high-dimensional UDIPs, the majority of the loci identified by JAGWAS were missed by previous IDP GWAS. This includes not only the original big40 GWAS but also subsequent brain imaging studies in the UK Biobank and other datasets^5,10,19^. Despite the significant advancement in loci discovery, using UDIPs as phenotypes presents an inevitable challenge caused by the difficulty in interpretation. In traditional IDP GWAS, each brain morphology measure can have a specific meaning, and regional analysis can be conducted to identify genetic variants with localized effects. However, with UDIPs, it is challenging to interpret what each UDIP represents since they are essentially interchangeable. Another limitation of JAGWAS is shared by all multivariate methods, i.e., multi-phenotype associations might be less intuitive to interpret compared to single-phenotype associations.

Such associations can usually be interpreted as a genetic variant being associated with at least one of the phenotypes, but multivariate methods do not directly identify which one or more phenotypes are associated.

The UDIPs are high-dimensional with moderate correlation, and each individual dimension is expected to be orderless and interchangeable, thus making it a naturally good fit for efficient multi-phenotype GWAS methods like JAGWAS. For highly correlated phenotypes, however, the correlation matrix might be close to singular and non-invertible. In that case, a Moore-Penrose generalized inverse of the correlation matrix should be used in JAGWAS. On the other hand, like many phenotypes, the genetic discovery of traditional brain IDPs is limited. Therefore, the primary challenge of the field is likely to transition into designing novel and robust descriptors of the brain morphology, in combination with efficient multivariate approaches to further enhance the genetic findings of imaging phenotypes. Our work represents an attempt in this direction and more future works are warranted. For example, multi-phenotype fine-mapping techniques have demonstrated the ability to identify putative causal variants by leveraging the shared structure across multiple phenotypes^63^. A future direction would be to explore memory-efficient multi-phenotype fine-mapping approaches for high-dimensional imaging phenotypes.

In conclusion, our work is a multi-phenotype GWAS application of UDIPs derived from an unsupervised deep learning approach. It offers a promising strategy for enhancing loci discovery in imaging phenotypes compared to the single-phenotype GWAS using minP approach. With the ultra-efficient multivariate approach JAGWAS, this method paves the way for future research to unravel intricate genotype-phenotype relationships, especially within the field of brain imaging.

## Methods

### Sample and dataset descriptions

We made use of UKB data obtained from the data repository under the project 24247. Subjects with White British ancestry were selected for this study. A total of 35,239 T1 and 34,145 T2 subjects were divided into discovery (22,880 T1/T2) and replication cohorts (T1=12,359/T2=11,265), subjects in replication cohort that are closely related to subjects in discovery cohort have been removed^15^. A total of 46,099 T1 (44,181 subjects) and 45,294 T2 (43,381 subjects) UKB MRIs (field ID 20252/field ID 20253) were downloaded on October 15, 2021. The 128-dimensional UDIPs, derived from an unsupervised deep learning model, are independent and exhibit higher heritability compared to traditional IDPs. They can also capture information from multiple regions of the brain. The details of the deep learning architecture and training can be found in Patel et al^15^.

### Single-phenotype GWAS procedure

The UKB v3 imputed dataset, following the comprehensive quality control procedures detailed by the UKB genetics team, was used for the single-phenotype GWAS. We additionally carried out standard quality check procedures, including removing SNPs with more than 5% missingness. We further set a minor allele frequency (MAF) threshold to remove low-frequency and rare variants with MAF < 0.01. Each of the 128 UDIPs (T1/T2) was pre-residualized through a linear model, with age (field ID 21003), age^2^, sex (field ID 31), sex x age, sex x age^2^, top 10 genetic PCs (field ID 22009), head size (field ID 25000), head position in scanner (field ID 25756-25758), scanner table position (field ID 25759), location of the assessment center (field ID 54) and date of attending assessment center (field ID 53) as covariates. We performed genome-wide scans on each of the pre-residualized and normalized UDIPs for 8,126,192 genetic variants, on the discovery, replication and meta-analysis cohort for both T1 and T2. We used fastGWA from GCTA (Genome-wide Complex Trait Analysis)^64^ (Version 1.94.1) package for running GWAS using 256 UDIPs obtained from T1(128 dimensions) and T2(128 dimensions) MRI linear mixed model association analysis with a sparse kinship matrix provided by the UK Biobank (field ID 22011/field ID 22012). By employing a linear mixed model, JAGWAS can increase the sample size by including related individuals. GWAS was run for both the discovery and the replication cohorts separately. For calling any SNP-UDIP pair genome-wide significant, we used the minP approach on 128 single-phenotype UDIP p-values at the significance level of 5 × 10^−8^/128 for the discovery cohort. The resulting summary statistics files were then passed onto JAGWAS for joint analysis. Details of the single-phenotype GWAS can be found in Patel et^20^.

### JAGWAS method

A multivariate association test statistic was derived for JAGWAS. Let *z_ij_* be the test statistic (z-score) obtained from the single-phenotype GWAS between the ith phenotype and jth SNP. Then let 𝒛_𝒋_ = [𝑧_1𝑗_, …, 𝑧_𝑘𝑗_] be the vector of z-scores across the 𝑘 phenotypes on the jth SNP. The JAGWAS method is based on a multivariate test statistic 𝑇_𝑗_ = 𝒛_𝒋_^𝑻^𝑹^−1^𝒛_𝒋_, where 𝑹 represents the phenotypic correlation matrix. Under the null hypothesis, this statistic asymptotically follows a chi-square distribution with k degrees of freedom. Therefore JAGWAS performs a chi-square test using the test statistic 𝒛_𝒋_^𝑻^𝑹^−1^𝒛_𝒋_, where 𝑹 is estimated by the k-by-k observed correlation matrix of scaled residuals. The scaled residuals are residuals obtained through a linear mixed model, divided by the residual variance estimate.

### Loci clumping

We adhered to the methodology outlined in our previous single-phenotype GWAS for loci clumping^15^, employing a two-step pruning process. Initially, significant SNPs were pruned at LD r^2^ = 0.6 to obtain a list of independent significant SNPs. Subsequently, these independent significant SNPs underwent further pruning at LD r^2^ = 0.1 to identify the independent lead SNPs. A genomic locus was defined as the smallest contiguous region encompassing all SNPs (including both GWAS markers and markers from the 1000 Genomes reference panel meeting the MAF threshold) with an r^2^ value exceeding 0.1 with the lead SNPs. Loci with adjacent physical distances less than 250 kb were merged together. Consequently, the presence of more than one lead SNP per locus is plausible. All loci clumping analyses were conducted using FUMA^29^.

### Meta-analysis

We used METAL(generic-metal-2011-03-25)^65^ to perform an inverse-variance-weighted fixed effect meta-analysis of single-phenotype GWAS summary statistics from the discovery and replication cohorts. The GWAS summary statistics files from the meta-analysis were then jointly analyzed using JAGWAS, with the covariance matrix computed from the weighted average of discovery and replication covariance matrices:

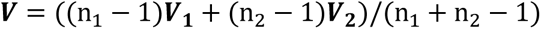

Where n_1_ and n_2_ denote the sample size of the discovery and replication cohort, 𝑽**_1_** and 𝑽**_2_** denote the covariance matrix of the discovery and replication cohort representatively. It has been shown that this pooled covariance matrix is a good estimate of the covariance matrix for meta-analysis^66^. The meta-analysis correlation matrix is then computed from this pooled covariance matrix 𝑽.

### Clustering analysis

Hierarchical clustering with the complete linkage method, which minimizes the maximum within-cluster distance, was applied to the vectors of optimal weights on the 128 UDIPs for each lead SNP. For any given weight vector 𝒘_𝒋_ with length 128 for SNP 𝑗, the univariate test statistic for a linear combination hypothesis 𝐻_0_ ∶ 𝒘_𝒋_^𝑻^𝜷_𝒋_ = 0 is: 𝒛_𝒋_^𝑻^𝒘_𝒋_(𝒘_𝒋_^𝑻^𝑹𝒘_𝒋_)^−1^𝒘_𝒋_^𝑻^𝒛_𝒋_. The optimal weight vector 𝒘_𝒋_ = 𝑹^−1^𝒛_𝒋_ maximizes this test statistic and gives the smallest p-value among all possible linear combinations (Supplementary information: Derivation of the optimal weights). The scale-free cosine distance metric was used to compute the similarity of optimal weight vectors between SNPs, and the cosine similarity matrix was used to indicate the color depth in the heatmap. The Python seaborn clustering package was used to derive the clusters and dendrograms^67^. Details of the derivation of optimal weights and the clustering procedure are available in the Supplementary information.

### Visualizing SNP effect

We visualized each lead SNP by generating a t-map of the MRI, comparing homozygous carriers to non-carriers in the UK Biobank dataset. For each voxel, the t-statistic was calculated as the difference in mean MRI signal intensity between the two groups, normalized by the pooled variance and sample size (Supplementary Fig. 7).

### Querying GWAS Catalog

We used FUMA to query candidate SNPs in each locus in GWAS to identify previously reported associations. FUMA directly calls GWAS catalog API and thus our results reflect the GWAS catalog as of May 2024. Candidate SNPs are defined by having r^2^ >= 0.6 with independent significant SNPs (See Methods: Loci clumping) not only in the GWAS variants but also in the 1000 Genomes reference panel. We filtered GWAS catalog results to include those with p-value < 5 × 10^-8^. We identified brain-related traits from the GWAS catalog results using careful manual inspection.

## Data availability

The GWAS summary statistics generated in this study have been uploaded to the GWAS Catalog https://www.ebi.ac.uk/gwas/ with study accession ID GCST90455631, GCST90455632, GCST90455633, and GCST90455634. The UDIPs are being uploaded to http://deependo.org.

## Code availability

The code is available via https://github.com/hanchenlab/JAGWAS (GPLv3 license).

## Data Availability

All data produced are available online (see Data and Code availability)

## Acknowledgments

This work was supported by grants from the National Institute on Aging (U01 AG070112).

## Ethics oversight

Our analysis was approved by the UTHealth Houston committee for the protection of human subjects under No. HSC-SBMI-20-1323. UKB has secured informed consent from the participants in the use of their data for approved research projects. UKB data was accessed via approved project 24247.

## References

1. Hariri, A. R. & Weinberger, D. R. Imaging genomics. Br. Med. Bull. 65, 259–270 (2003).

2. Hariri, A. R., Drabant, E. M. & Weinberger, D. R. Imaging genetics: perspectives from studies of genetically driven variation in serotonin function and corticolimbic affective processing. Biol. Psychiatry 59, 888–897 (2006).

3. Meyer-Lindenberg, A. The future of fMRI and genetics research. Neuroimage 62, 1286– 1292 (2012).

4. Miller, K. L. et al. Multimodal population brain imaging in the UK Biobank prospective epidemiological study. Nat. Neurosci. 19, 1523–1536 (2016).

5. Elliott, L. T. et al. Genome-wide association studies of brain imaging phenotypes in UK Biobank. Nature 562, 210–216 (2018).

6. Gong, W., Beckmann, C. F. & Smith, S. M. Phenotype discovery from population brain imaging. Med. Image Anal. 71, 102050 (2021).

7. Gong, W., Bai, S., Zheng, Y. Q., Smith, S. M. & Beckmann, C. F. Supervised Phenotype Discovery From Multimodal Brain Imaging. IEEE Trans. Med. Imaging 42, 834–849 (2023).

8. Uffelmann, E. et al. Genome-wide association studies. Nat. Rev. Methods Primers 1, 59 (2021).

9. Sudlow, C. et al. UK biobank: an open access resource for identifying the causes of a wide range of complex diseases of middle and old age. PLoS Med. 12, e1001779 (2015).

10. Smith, S. M. et al. An expanded set of genome-wide association studies of brain imaging phenotypes in UK Biobank. Nat. Neurosci. 24, 737–745 (2021).

11. Shadrin, A. A. et al. Vertex-wise multivariate genome-wide association study identifies 780 unique genetic loci associated with cortical morphology. Neuroimage 244, 118603 (2021).

12. Zhao, B. et al. Genome-wide association analysis of 19,629 individuals identifies variants influencing regional brain volumes and refines their genetic co-architecture with cognitive and mental health traits. Nat. Genet. 51, 1637–1644 (2019).

13. Alfaro-Almagro, F. et al. Image processing and Quality Control for the first 10,000 brain imaging datasets from UK Biobank. Neuroimage 166, 400–424 (2018).

14. Sweitzer, M. M., Donny, E. C. & Hariri, A. R. Imaging genetics and the neurobiological basis of individual differences in vulnerability to addiction. Drug Alcohol Depend. 123 **Suppl 1**, S59–71 (2012).

15. Patel, K. et al. Unsupervised deep representation learning enables phenotype discovery for genetic association studies of brain imaging. *Commun*. Biol. 7, 414 (2024).

16. Hibar, D. P. et al. Novel genetic loci associated with hippocampal volume. Nat. Commun. 8, 13624 (2017).

17. van der Lee, S. J. et al. A genome-wide association study identifies genetic loci associated with specific lobar brain volumes. *Commun*. Biol. 2, 285 (2019).

18. Grasby, K. L. et al. The genetic architecture of the human cerebral cortex. Science 367, eaay6690 (2020).

19. van der Meer, D. et al. Understanding the genetic determinants of the brain with MOSTest. Nat. Commun. 11, 3512 (2020).

20. Hawrylycz, M. et al. Canonical genetic signatures of the adult human brain. Nat. Neurosci. 18, 1832–1844 (2015).

21. Park, S. et al. Multivariate genomic analysis of 5 million people elucidates the genetic architecture of shared components of the metabolic syndrome. Nat. Genet. (2024) doi:10.1038/s41588-024-01933-1.

22. O’Reilly, P. F. et al. MultiPhen: joint model of multiple phenotypes can increase discovery in GWAS. PLoS One 7, e34861 (2012).

23. Ferreira, M. A. & Purcell, S. M. A multivariate test of association. Bioinformatics 25, 132– 133 (2009).

24. Shen, X. et al. Multivariate discovery and replication of five novel loci associated with Immunoglobulin G N-glycosylation. Nat. Commun. 8, 447 (2017).

25. Ray, D. & Boehnke, M. Methods for meta-analysis of multiple traits using GWAS summary statistics. Genet. Epidemiol. 42, 134–145 (2018).

26. Longford, N. T. Multivariate Analysis of Variance. in International Encyclopedia of Education (Third Edition) (ed. Barry, P. P. B. E.) 319–323 (Elsevier, Oxford, 2010).

27. Turley, P. et al. Multi-trait analysis of genome-wide association summary statistics using MTAG. Nat. Genet. 50, 229–237 (2018).

28. Sollis, E. et al. The NHGRI-EBI GWAS Catalog: knowledgebase and deposition resource. Nucleic Acids Res. 51, D977–D985 (2023).

29. Watanabe, K., Taskesen, E., van Bochoven, A. & Posthuma, D. Functional mapping and annotation of genetic associations with FUMA. Nat. Commun. 8, 1826 (2017).

30. Park, K. W. et al. Ethnicity- and sex-specific genome wide association study on Parkinson’s disease. NPJ Parkinsons Dis. 9, 141 (2023).

31. Wang, H., Yi, Z. & Shi, T. Novel loci and potential mechanisms of major depressive disorder, bipolar disorder, and schizophrenia. Sci. China Life Sci. 65, 167–183 (2022).

32. Mekki, Y. et al. The genetic architecture of language functional connectivity. Neuroimage 249, 118795 (2022).

33. Watanabe, K. et al. Genome-wide meta-analysis of insomnia prioritizes genes associated with metabolic and psychiatric pathways. Nat. Genet. 54, 1125–1132 (2022).

34. Hofer, E. et al. Genetic correlations and genome-wide associations of cortical structure in general population samples of 22,824 adults. Nat. Commun. 11, 4796 (2020).

35. van der Meer, D. et al. The genetic architecture of human cortical folding. Sci. Adv. 7, eabj9446 (2021).

36. Liu, N. et al. Cross-ancestry genome-wide association meta-analyses of hippocampal and subfield volumes. Nat. Genet. 55, 1126–1137 (2023).

37. Khlif, M. S. et al. Hippocampal subfield volumes are associated with verbal memory after first-ever ischemic stroke. Alzheimers Dement. (Amst.) 13, e12195 (2021).

38. Zhang, H.-L. et al. HDAC9-mediated calmodulin deacetylation induces memory impairment in Alzheimer’s disease. CNS Neurosci. Ther. 30, e14573 (2024).

39. Ji, Y. et al. Exploring functional dysconnectivity in schizophrenia: alterations in eigenvector centrality mapping and insights into related genes from transcriptional profiles. Schizophrenia (Heidelb.) 10, 37 (2024).

40. Rahman, M. A. & Liu, J. A genome-wide association study coupled with machine learning approaches to identify influential demographic and genomic factors underlying Parkinson’s disease. Front. Genet. 14, 1230579 (2023).

41. Blokland, G. A. M. et al. Sex-dependent shared and nonshared genetic architecture across mood and psychotic disorders. Biol. Psychiatry 91, 102–117 (2022).

42. Mendoza, J. E. Lingual Gyrus. in Encyclopedia of Clinical Neuropsychology 1–1 (Springer International Publishing, Cham, 2017).

43. Galetta, S. L. Occipital Lobe. in Encyclopedia of the Neurological Sciences (eds. Aminoff, M. J. & Daroff, R. B.) 626–632 (Elsevier, 2014).

44. Gagliano Taliun, S. A., et al. Exploring and visualizing large-scale genetic associations by using PheWeb. Nat. Genet. 52, 550–552 (2020).

45. Liu, X. et al. Decreased functional connectivity between the dorsal anterior cingulate cortex and lingual gyrus in Alzheimer’s disease patients with depression. Behav. Brain Res. 326, 132–138 (2017).

46. Tohid, H., Faizan, M. & Faizan, U. Alterations of the occipital lobe in schizophrenia. Neurosciences 20, 213–224 (2015).

47. Adewuyi, E. O., O’Brien, E. K., Nyholt, D. R., Porter, T. & Laws, S. M. A large-scale genome-wide cross-trait analysis reveals shared genetic architecture between Alzheimer’s disease and gastrointestinal tract disorders. *Commun*. Biol. 5, 691 (2022).

48. Urbán, N. & Guillemot, F. Neurogenesis in the embryonic and adult brain: same regulators, different roles. Front. Cell. Neurosci. 8, 396 (2014).

49. Vieira, M. S. et al. Neural stem cell differentiation into mature neurons: Mechanisms of regulation and biotechnological applications. Biotechnol. Adv. 36, 1946–1970 (2018).

50. Furlanis, E. & Scheiffele, P. Regulation of neuronal differentiation, function, and plasticity by alternative splicing. Annu. Rev. Cell Dev. Biol. 34, 451–469 (2018).

51. Tyng, C. M., Amin, H. U., Saad, M. N. M. & Malik, A. S. The influences of emotion on learning and memory. Front. Psychol. 8, 1454 (2017).

52. de Jong, L. W. et al. Strongly reduced volumes of putamen and thalamus in Alzheimer’s disease: an MRI study. Brain 131, 3277–3285 (2008).

53. Burdakov, D. & Peleg-Raibstein, D. The hypothalamus as a primary coordinator of memory updating. Physiol. Behav. 223, 112988 (2020).

54. Foerde, K. & Shohamy, D. The role of the basal ganglia in learning and memory: insight from Parkinson’s disease. Neurobiol. Learn. Mem. 96, 624–636 (2011).

55. Wang, H. et al. Genome-wide interaction analysis of pathological hallmarks in Alzheimer’s disease. Neurobiol. Aging 93, 61–68 (2020).

56. Gouveia, C. et al. Genome-wide association of polygenic risk extremes for Alzheimer’s disease in the UK Biobank. Sci. Rep. 12, 8404 (2022).

57. Rao, S., Baranova, A., Yao, Y., Wang, J. & Zhang, F. Genetic relationships between attention-deficit/hyperactivity disorder, autism spectrum disorder, and intelligence. Neuropsychobiology 81, 484–496 (2022).

58. Lam, M. et al. Comparative genetic architectures of schizophrenia in East Asian and European populations. Nat. Genet. 51, 1670–1678 (2019).

59. Goes, F. S. et al. Genome-wide association study of schizophrenia in Ashkenazi Jews. Am. J. Med. Genet. B Neuropsychiatr. Genet. 168, 649–659 (2015).

60. Sherva, R. et al. Genome-wide association study of the rate of cognitive decline in Alzheimer’s disease. Alzheimers. Dement. 10, 45–52 (2014).

61. Schwartzentruber, J. et al. Genome-wide meta-analysis, fine-mapping and integrative prioritization implicate new Alzheimer’s disease risk genes. Nat. Genet. 53, 392–402 (2021).

62. Balestri, W. et al. Modeling the neuroimmune system in Alzheimer’s and Parkinson’s diseases. J. Neuroinflammation 21, 32 (2024).

63. Zou, Y., Carbonetto, P., Xie, D., Wang, G. & Stephens, M. Fast and flexible joint fine-mapping of multiple traits via the Sum of Single Effects model. bioRxivorg (2024) doi:10.1101/2023.04.14.536893.

64. Jiang, L. et al. A resource-efficient tool for mixed model association analysis of large-scale data. Nat. Genet. 51, 1749–1755 (2019).

65. Willer, C. J., Li, Y. & Abecasis, G. R. METAL: fast and efficient meta-analysis of genomewide association scans. Bioinformatics 26, 2190–2191 (2010).

66. Lee, J. A., Dobbin, K. K. & Ahn, J. Covariance adjustment for batch effect in gene expression data. Stat. Med. 33, 2681–2695 (2014).

67. Waskom, M. L. seaborn: statistical data visualization. J. Open Source Softw. 6(60), 3021 (2021).

