## Supplementary Information for "Efficient multi-phenotype genome-wide analysis identifies genetic associations for unsupervised deep-learning-derived high-dimensional brain imaging phenotypes"

### 1. Supplementary Figures 2. Supplementary Notes

### Supplementary Figures

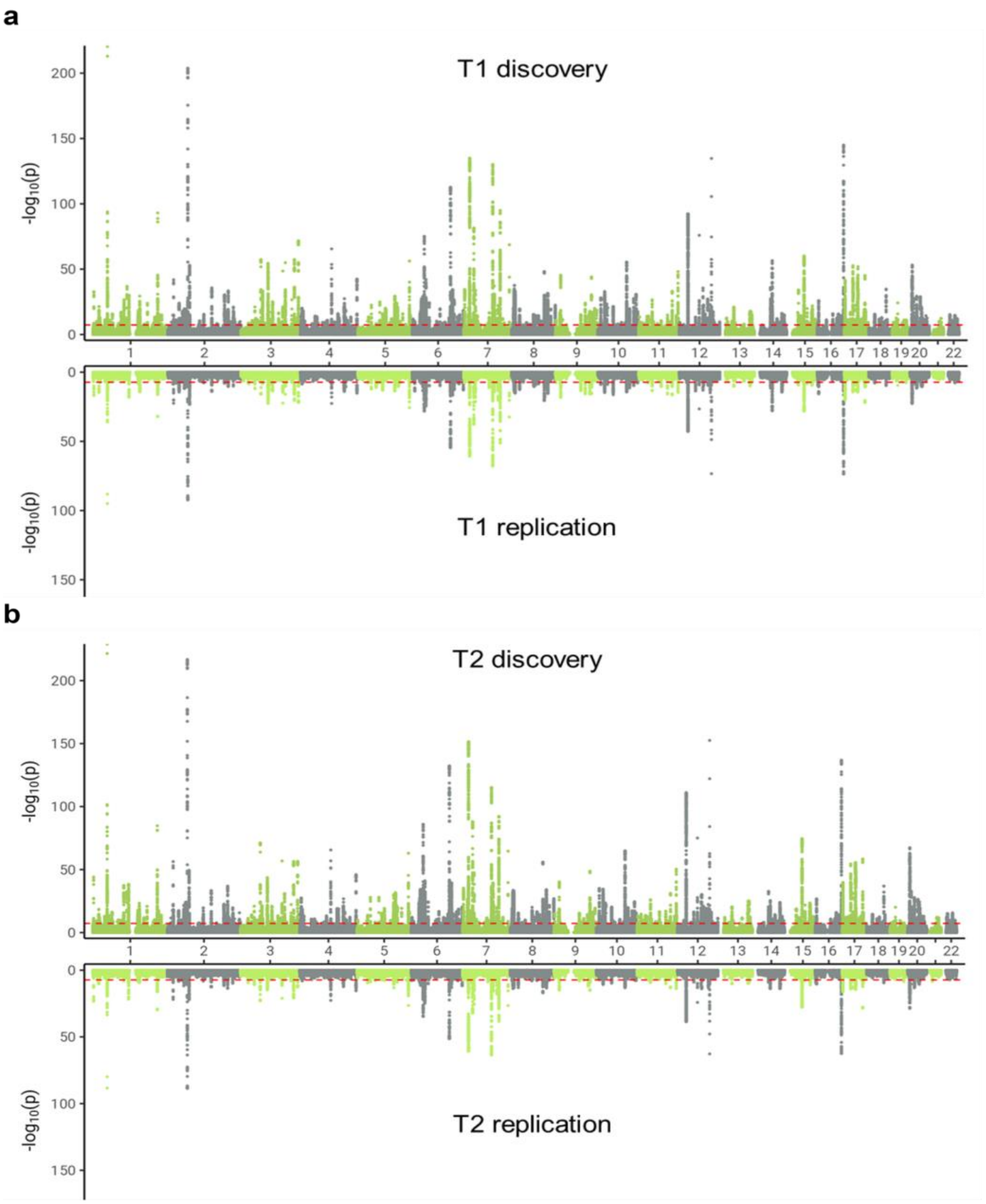


**Supplementary Fig 1. Miami plots of GWAS results from discovery and replication cohorts.** Miami plots of -log10 p-values from the JAGWAS results of discovery cohort vs. replication cohort for T1 (a) and T2 (b) respectively. The y axis represents the -log10 p-value and x axis shows the relative genomic location, grouped by chromosome, and the red dashed lines indicate the genome-wide significance threshold of $5 \times 10^{-8}$.

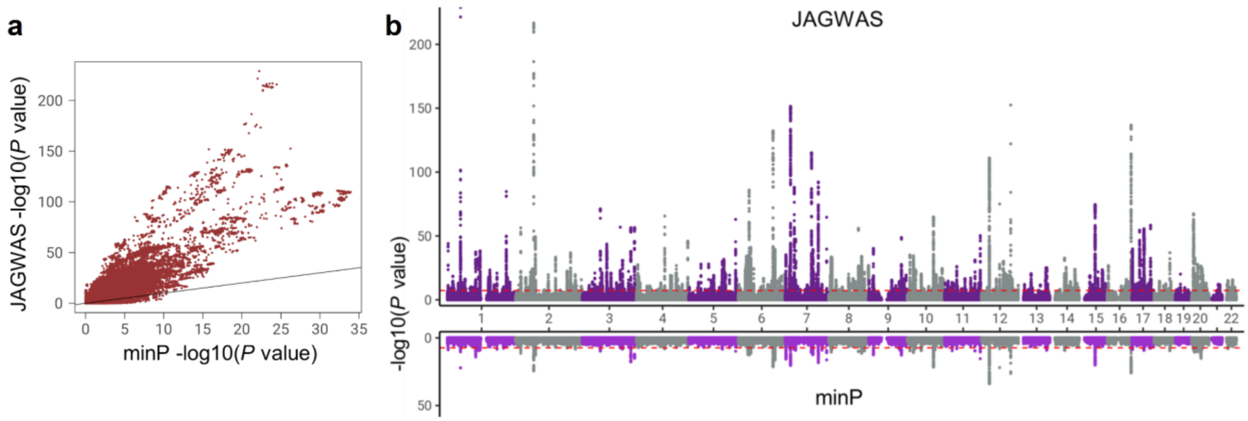


**Supplementary Fig. 2 Scatter and Miami plot of GWAS results of JAGWAS vs. minP for T2 (discovery cohort). a**. Scatter plot of -log10 p-values from JAGWAS vs. minP. **b**. Miami plot of -log10 p-values of JAGWAS vs. minP, with JAGWAS on the top half and minP on the bottom half. The y axis represents the -log10 p-value and x axis shows the relative genomic location, grouped by chromosome, and the red dashed lines indicate the genome-wide significance threshold of $5 \times 10^{-8}$.

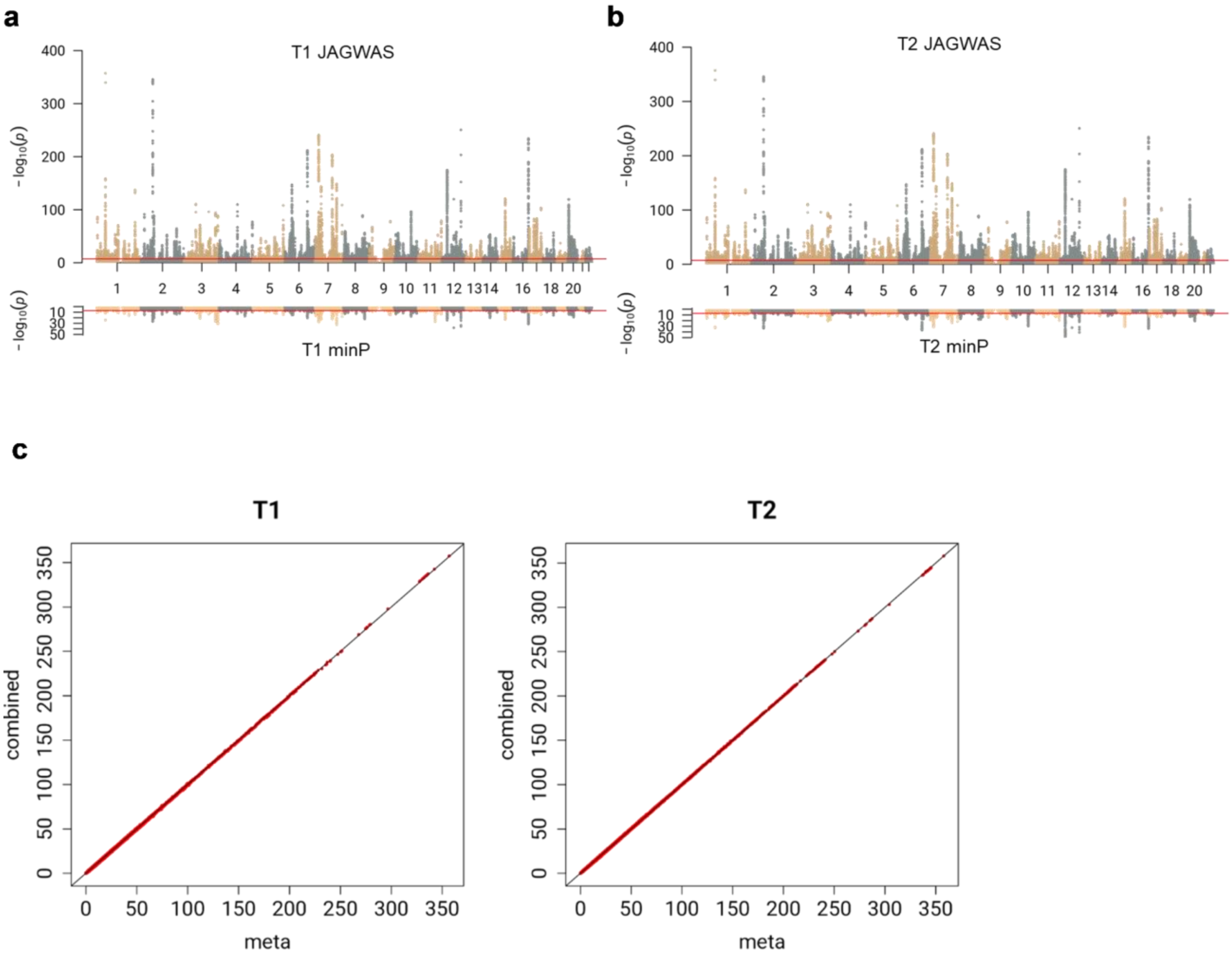


**Supplementary Fig. 3 Results from meta-analysis of discovery and replication cohorts. a-b.** Miami plots of -log10 p-values from the JAGWAS results of meta-analysis of discovery and replication cohort, for T1 (a) and T2 (b) respectively. The y axis represents the -log10 p-value and x axis shows the relative genomic location, grouped by chromosome, and the red dashed lines indicate the genome-wide significance threshold of $5 \times 10^{-8}$. **c.** Scatter plot of -log10 p-values from JAGWAS results in the combined sample on y axis (discovery plus replication cohort) vs. in the meta-analysis on x axis.

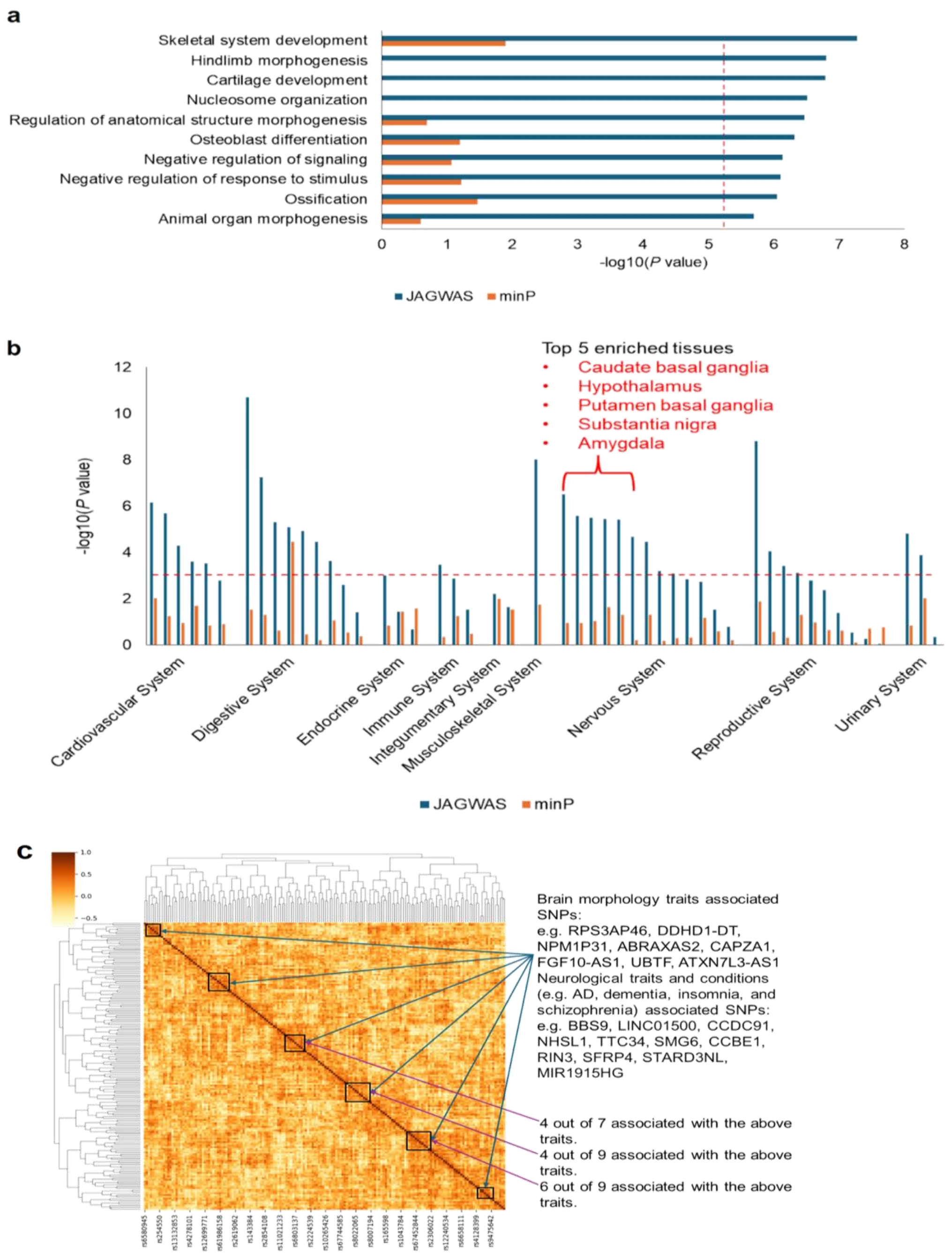


**Supplementary Fig. 4 Enrichment and clustering analyses indicate high neurobiological relevance of JAGWAS hits (T2). a.** Results from gene-set enrichment analyses for JAGWAS and minP. The top 10 most significant Biological Process Gene Ontology sets for JAGWAS (blue bars) are shown on the y axis, -log10 p-value on the x axis. The corresponding minP p-values are represented by the orange bars. The red dashed significance line (0.05/6500) has been adjusted using a Bonferroni correction. **b.** Results from tissue enrichment analyses for for JAGWAS (blue bars) and minP (orange bars). The GTEx v8 54 tissue types classified by organ systems are shown on the x axis, -log10 two-side DEG p-value on the y axis. JAGWAS and minP p-values are represented by blue and orange bars respectively. The red dashed significance line (0.05/54) has been adjusted using a Bonferroni correction. **c.** Clustering lead SNPs by their vectors of optimal weights on the 128 UDIPs, in order to group SNPs by similar pleiotropic effects. Darker colors indicate highly similar SNPs based on cosine similarity.

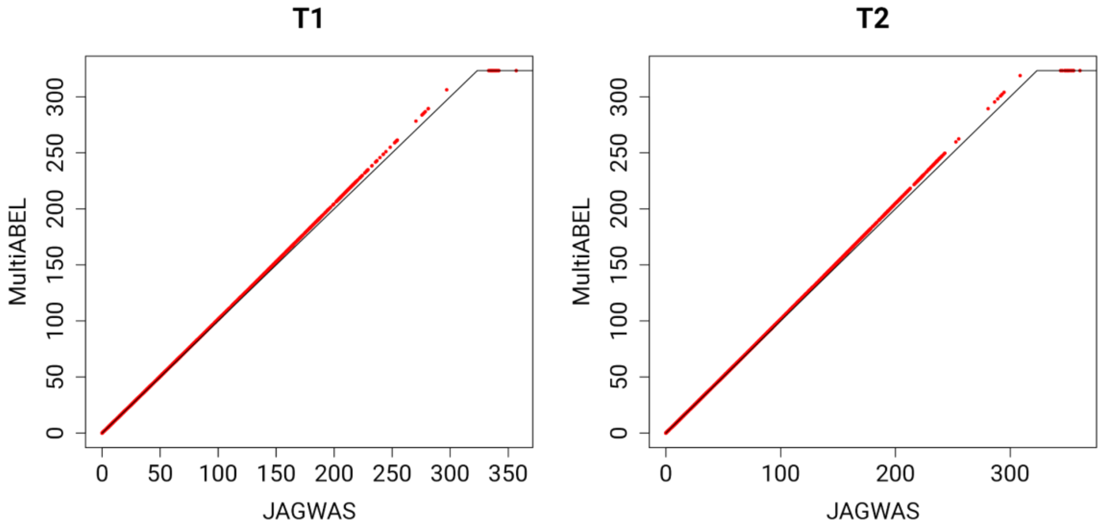


**Supplementary Fig. 5 Scatter plot of JAGWAS vs. MultiABEL.** Scatter plot of -log10 p-values from JAGWAS results using the meta-analysis sample vs. -log10 p-values from MultiABEL results using the same sample. MultiABEL p-values are on the y axis while JAGWAS p-values are on the x axis. Note that the plot is clipped at -log10($5 \times 10^{-324}$) = 323.30 for MultiABEL p-values since this is the smallest p-value R can output, while JAGWAS can output p-values on -log10 scale thus avoiding such a lower limit.

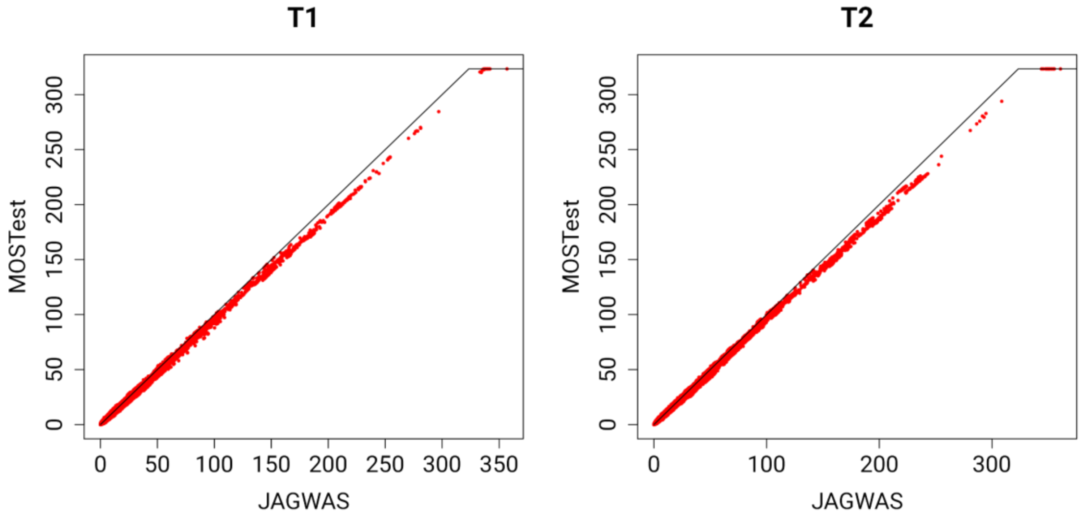


**Supplementary Fig. 6 Scatter plots of JAGWAS vs. MOSTest.** Scatter plots of -log10 p-values from JAGWAS results using the combined sample vs. -log10 p-values from MOSTest results using an unrelated subset of the combined sample. The MOSTest -log10 p-values are on the y axis while JAGWAS p-values are on the x axis. Note that the plot is clipped at -log10($5 \times 10^{-324}$) = 323.30 for MOSTest p-values since this is the smallest p-value MOSTest can output.

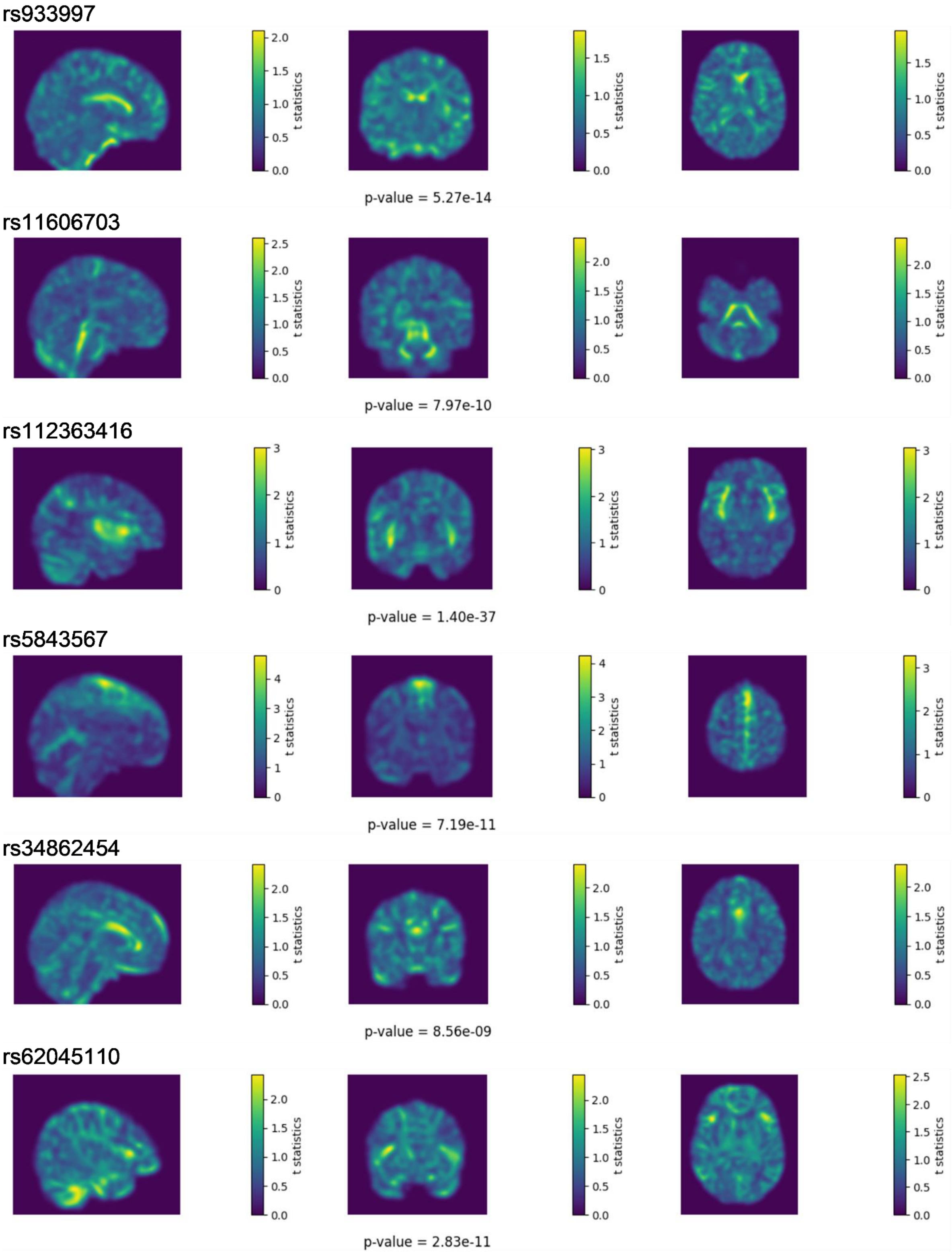


**Supplementary Fig. 7** Visualization of the effect of SNPs showing localized patterns of differences between homozygous carriers and non-carriers.

### Supplementary Notes

#### Derivation of the optimal weights on the 128 UDIPs

Recall that the JAGWAS test statistics $z^T R^{-1} z$ is testing the null hypothesis:

$$H_0 : \boldsymbol{\beta} = 0$$

where $\boldsymbol{\beta}$ represents a length 128 vector of genetic effects of a single SNP on the UDIPs.
The univariate test statistic for a linear combination hypothesis $H_0 : \boldsymbol{w^T\beta} = 0$ is:

$$f(\boldsymbol{w}) = (\boldsymbol{w^T z})^2 (\boldsymbol{w^T R w})^{-1}$$

as a function on a length 128 vector $\boldsymbol{w}$. Under the null hypothesis, it follows a chi-square distribution with 1 degree of freedom.
Maximizing this univariate test statistic would result in the smallest p-value, with $\boldsymbol{w}$ representing the best linear combination for a univariate test in the high-dimensional space. Both the length 128 vector of z-scores $\boldsymbol{z}$ and the 128 by 128 correlation matrix $\boldsymbol{R}$ are known from the data.
The first derivative of $f(\boldsymbol{w})$:

$$\frac{\partial \boldsymbol{f}}{\partial \boldsymbol{w}} = 2(\boldsymbol{w^T R w})^{-1}(\boldsymbol{z z^T w}) - 2(\boldsymbol{w^T R w})^{-2}(\boldsymbol{w^T z})^2 \boldsymbol{R w} = 0$$

$$(\boldsymbol{w^T R w})^{-1}(\boldsymbol{z z^T w}) = (\boldsymbol{w^T R w})^{-2}(\boldsymbol{w^T z})^2 \boldsymbol{R w}$$

$$(\boldsymbol{w^T R w})\boldsymbol{z} = (\boldsymbol{z^T w})\boldsymbol{R w}$$

And $\boldsymbol{w} = \boldsymbol{R}^{-1}\boldsymbol{z}$ is one solution for the above equation.
The second derivative of $f(\boldsymbol{w})$:

$$\frac{\partial^2 \boldsymbol{f}}{\partial \boldsymbol{w} \partial \boldsymbol{w^T}} = -4(\boldsymbol{w^T R w})^{-2}(\boldsymbol{z z^T w})\boldsymbol{w^T R} + 2(\boldsymbol{w^T R w})^{-1}\boldsymbol{z z^T} +$$

$$8(\boldsymbol{w^T R w})^{-3}(\boldsymbol{w^T z})^2 \boldsymbol{R w w^T R} - 4(\boldsymbol{w^T R w})^{-2}(\boldsymbol{w^T z})\boldsymbol{R w z^T} - 2(\boldsymbol{w^T R w})^{-2}(\boldsymbol{w^T z})^2 \boldsymbol{R}$$

$$\frac{\partial^2 \boldsymbol{f}}{\partial \boldsymbol{w} \partial \boldsymbol{w^T}}\Big|_{\boldsymbol{w}=\boldsymbol{R}^{-1}\boldsymbol{z}} = -4(\boldsymbol{z^T R}^{-1}\boldsymbol{z})^{-1}\boldsymbol{z z^T} + 2(\boldsymbol{z^T R}^{-1}\boldsymbol{z})^{-1}\boldsymbol{z z^T} + 8(\boldsymbol{z^T R}^{-1}\boldsymbol{z})^{-1}\boldsymbol{z z^T}$$

$$-4(\boldsymbol{z^T R}^{-1}\boldsymbol{z})^{-1}\boldsymbol{z z^T} - 2\boldsymbol{R}$$

$$= 2(\boldsymbol{z^T R}^{-1}\boldsymbol{z})^{-1}\boldsymbol{z z^T} - 2\boldsymbol{R} = -2(\boldsymbol{R}^{\frac{1}{2}}\left(\boldsymbol{I} - \boldsymbol{R}^{-\frac{1}{2}}\boldsymbol{z}(\boldsymbol{z^T R}^{-1}\boldsymbol{z})^{-1}\boldsymbol{z^T R}^{-\frac{1}{2}}\right)\boldsymbol{R}^{\frac{1}{2}})$$

The matrix $\left(\boldsymbol{I} - \boldsymbol{R}^{-\frac{1}{2}}\boldsymbol{z}(\boldsymbol{z^T R}^{-1}\boldsymbol{z})^{-1}\boldsymbol{z^T R}^{-\frac{1}{2}}\right)$ is idempotent and therefore is positive semi-definite.
Consequently, $-2(\boldsymbol{R}^{\frac{1}{2}}\left(\boldsymbol{I} - \boldsymbol{R}^{-\frac{1}{2}}\boldsymbol{z}(\boldsymbol{z^T R}^{-1}\boldsymbol{z})^{-1}\boldsymbol{z^T R}^{-\frac{1}{2}}\right)\boldsymbol{R}^{\frac{1}{2}})$ is symmetric and negative semi-definite.
Thence, $\boldsymbol{w} = \boldsymbol{R}^{-1}\boldsymbol{z}$ maximizes the univariate test statistic for a linear combination hypothesis $H_0 : \boldsymbol{w^T \beta} = 0$.

#### Clustering SNPs based on their vectors of optimal weights

The scale-free cosine distance metric was used to compute the similarity between each lead SNP's vector of optimal weights. Cosine similarity was chosen due to its ability to measure the directional alignment between vectors, irrespective of the magnitudes of weights from different dimensions.
Given optimal weights $\boldsymbol{w_i}$ and $\boldsymbol{w_j}$ of two SNPs, cosine similarity measures the cosine of the angle $\theta_{ij}$ between $\boldsymbol{w_i}$ and $\boldsymbol{w_j}$:

$$cos(\theta_{ij}) = \frac{\boldsymbol{w_i}^{\boldsymbol{T}}\boldsymbol{w_j}}{\sqrt{\boldsymbol{w_i}^{\boldsymbol{T}}\boldsymbol{w_i}}\sqrt{\boldsymbol{w_j}^{\boldsymbol{T}}\boldsymbol{w_j}}}$$

By focusing on the angle between vectors rather than their length, cosine similarity effectively captures relationships where SNPs exhibit similar patterns of association across phenotypes, even if the strength of those associations differs. A hierarchical clustering heatmap was generated based on the cosine similarity matrix of the lead SNPs, to identify SNPs with similar pleiotropic effects.
